# Critical Complications of COVID-19: A systematic Review and Meta-Analysis study

**DOI:** 10.1101/2020.06.14.20130955

**Authors:** Kimia Vakili, Mobina Fathi, Fatemeh Sayehmiri, Ashraf Mohamadkhani, Mohammadreza Hajiesmaeili, Mostafa Rezaei-Tavirani, Aiyoub Pezeshgi

## Abstract

**Background:** Coronavirus disease 2019 (COVID-19) is a novel coronavirus infection that has spread worldwide in a short period and caused a pandemic. The goal of this meta-analysis is to evaluate the prevalence of most common symptoms and complications of COVID-19.

**Methods:** All related studies assessing the clinical complications of COVID-19 have been identified through web search databases (PubMed and Scopus). Relevant data were extracted from these studies and analyzed by stata (ver 14) random-effects model. The heterogeneity of studies were assessed by *I*^*2*^ index. The publication bias was examined by Funnel plots and Egger’s test.

**Results:** 30 studies were in our meta-analysis including 6 389 infected patients. The prevalence of most common symptoms were: fever 84.30% (95% CI: 77.13-90.37; I^2^=97.74%), cough 63.01% (95% CI: 57.63-68.23; I^2^=93.73%), dyspnea 37.16% (95% CI: 27.31-47.57%; I^2^=98.32%), fatigue 34.22% (95% CI: 26.29-42.62; I^2^=97.29%) and diarrhea 11.47 %(95% CI: 6.96-16.87; I^2^=95.58%), respectively. The most prevalent complications were acute respiratory distress syndrome (ARDS) 33.15% (95% CI: 23.35-43.73; I^2^=98.56%), acute cardiac injury 13.77% (95% CI: 9.66-18.45; I^2^=91.36%), arrhythmia 16.64% (95% CI: 9.34-25.5; I^2^=92.29%), heart failure 11.50% (95% CI: 3.45-22.83; I^2^=89.48%), and acute kidney injury (AKI) 8.40 %(95% CI: 5.15-12.31; I^2^=95.22%, respectively. According to our analysis, mortality rate of COVID-19 patients were 12.29% (95% CI: 6.20-19.99; I^2^=98.29%).

**Conclusion:** We assessed the prevalence of the main clinical complications of COVID-19 and found that after respiratory complications, cardiac and renal complications are the most common clinical complications of COVID-19.

**Highlights:**

▪ The most prevalent complication among critical cases of COVID-19 is ARDS.
▪ After pulmonary complications, cardiovascular complications (like arrhythmia, heart failure and acute cardiac injury) are the most important threats for COVID-19 patients.
▪ Renal complications (like AKI) happen as a result of COVID-19, but they are less prevalent than pulmonary and cardiovascular complications.

## 1. Introduction

Since the first emergence of a cluster of pneumonia cases in Wuhan, China on December 2019, the global attraction has been drawn to this disease. The China Center for Disease Control and Prevention, on 9 January 2020, stated that the cause of this newly emerged disease is a novel virus from coronavirideae family, which has been further named severe acute respiratory syndrome coronavirus 2 (SARS-CoV-2) and the disease caused by this virus was called corona virus disease 2019 (COVID-19). Unfortunately, this epidemic was not limited to China; the World Health Organization (WHO), on 11 March 2020, announced COVID-19 outbreak a pandemic (1, 2). According to WHO, until the time that we are writing this (11 May 2020), more than 4 000 000 individuals were diagnosed with COVID-19 and unfortunately about 279 000 of them have died (3). Based on the latest reports, the most common symptoms of COVID-19 are fever, dry cough, fatigue, dyspnea and diarrhea (4).

The main clinical complications of COVID-19 are related to respiratory system, from a simple pneumonia in mild cases to acute respiratory distress syndrome (ARDS) and shock in severe patients. The angiotensin-converting enzyme 2 (ACE2) is the host cell receptor responsible for the entry of SARS-CoV-2 and the facilitation of infection. Lung, heart, kidney and intestine cells express ACE2. According to animal models, ACE2 is downregulated in lung injuries (5); therefore, severe lung inflammation due to SARS-CoV-2 infection can lead to dysregulation of the renin-angiotensin system and further development of ARDS.

Furthermore, SARS-CoV-2 is expected to affect all tissues that express ACE2. Therefore, in addition to respiratory manifestations, cardiovascular and renal complications may occur following COVID-19 infection. In practice, growing evidence demonstrates cardiovascular involvement in COVID-19 disease and its negative impact on prognosis (6). There are also many studies which confirm that acute and chronic renal injuries are expected due to the high expression of ACE2 in the renal tubular epithelium (7).

Since these complications are extremely life-threatening, their prevalence in the clinical outcomes of patients can act as a determining factor in the morbidity and mortality rate of the disease. Therefore, this article aims to determine the prevalence of critical complications of patients with COVID-19 by systematically reviewing recent literatures and analyzing related data.

## 2. Method and material

### 2.1. Study selection

To find eligible cross-sectional and case-control studies published on COVID-19 until 1^st^ May 2020, we conducted a web search with citations in the PubMed. Two authors conducted the search process independently. Our search queries included: ‘ARDS AND COVID’, ‘((Heart) OR Cardio-) AND COVID’, ‘((Kidney) OR Renal) AND COVID’ and, ‘Complications AND COVID’. On the basis of paper titles, eligible studies were chosen. In addition, the reference lists of all related reviews (narrative and systematic) were searched to find more related articles.

### 2.2. Inclusion and exclusion criteria

All the studies in which the clinical complications of COVID-19 were analyzed were examined. To be included in the final analysis, screened studies must report data on the prevalence of each clinical complication in COVID-19 patients, including ARDS, acute heart damage, arrhythmia, heart failure and AKI. Only human studies have been included. The exclusion criteria in our search strategy were systematic reviews and meta-analyzes, duplicate publications, non-human studies, lack of sufficient data and non-English publications. Since COVID-19 is a newly emerged disease and the literature related to this disease is still limited, the quality of the included literatures has not been evaluated.

### 2.3. Data extraction

For all studies, related data including the name of the first author, date of publication, location of publication, sample size, sample age, sample gender, prevalence of symptoms (including: fever, cough, dyspnea, fatigue, and diarrhea), critical complications (including: ARDS, acute cardiac injury, arrhythmia, heart failure, and AKI) and clinical outcomes (mortality rate) were extracted. To avoid conflicting results, the authors extracted data from each study separately and then the results were compared.

### 2.4. Statistical analysis

The prevalence of cardiovascular and renal complications was the effect size in this study. Using the binomial distribution, its variance was assessed (with 95% confidence interval). Average weight was applied to combine the prevalence of different studies. The inverse relationship was existed between the study weight and its variance. Q statistics and I^2^ index with α significance level of less than 10% were applied to investigate the heterogeneity. Meta-analysis (random effects model) were applied, when the studies were heterogeneous. For data analysis, STATA software (version 14) was applied. The Metaprop (meta-analysis for proportion) command was applied in STATA and when p was close to 0 or 1. In order to stabilize the variances, we applied Freeman-Tukey Double Arcsine Transformation (8). In metaprop two variables in the format such that p = n/N are required to be declared. This study was carried out under the approval of ethics committee of Shahid Beheshti University of Medical Sciences (IR.SBMU.RETECH.REC.1399.083).

## 3. Result

### 3.1. Study Selection

For analysis, we used PRISMA checklist (9). Initially, 137 studies were identified through database searches, and 44 additional studies through other sources. Of these 181 studies, 61 of them were excluded due to duplication. After screening the abstracts, other 55 articles were excluded too. Therefore, the full-text of 85 remaining were evaluated and 28 other studies were excluded for some reasons (Lack of enough information). After careful review of these selected studies, 30 published articles between February 2020 and April 2020 were included for the further analysis (Fig. 1, Table 1).

**Table 1.** Baseline characteristics of studies included in this meta-analysis.

| Study (ref) | Place | Patients (No.) |  |  | Age (Median) | Symptoms (P.) |  |  |  |  | Complications (P.) |  |  | Mortality (%) |
| --- | --- | --- | --- | --- | --- | --- | --- | --- | --- | --- | --- | --- | --- | --- |
|  |  | All | Male | Female |  | Fever | Cough | Fatigue | Diarrhea | Dyspnea | Acute Cardiac Injury | AKI | ARDS |  |
| Fei Zhou et al. <sup>(30)</sup> | China | 191 | 119 | 72 | 56 | 94 | 79 | 23 | 5 | 29 | 17 | 15 | 54 | 28 |
| Yingzhen Du et al. <sup>(31)</sup> | China | 85 | 62 | 23 |  | 92 | 22 | 59 | 19 | 59 | 5 |  | 47 |  |
| Lang Wang et al. <sup>(32)</sup> | China | 339 | 166 | 173 | 69 | 92 | 53 | 40 | 13 | 41 | 21 | 8 | 21 | 19 |
| Xun Li et al. <sup>(33)</sup> | China | 25 | 10 | 15 | 73 |  |  |  |  | 92 |  |  | 92 | 100 |
| Nanshan Chen et al. <sup>(14)</sup> | China | 99 | 67 | 32 |  | 83 | 82 | 11 | 2 | 31 |  | 3 | 17 | 11 |
| Dawei Wang et al. <sup>(34)</sup> | China | 138 | 75 | 63 |  | 99 | 59 | 70 | 10 | 31 | 7 | 4 | 20 | 4 |
| Wei-jie Guan et al. <sup>(4)</sup> | China | 1099 | 637 | 459 | 47 | 44 | 68 | 38 | 4 | 19 |  | 0.5 | 3 | 1 |
| Xiaobo Yang et al. <sup>(35)</sup> | China | 52 | 35 | 17 |  | 98 | 77 | 12 |  | 64 |  | 30 | 67 | 62 |
| Tao Guo et al. <sup>(36)</sup> | China | 187 | 91 | 96 |  |  |  |  |  |  |  | 15 | 25 | 23 |
| Yingxia Liu et al. <sup>(37)</sup> | China | 12 | 8 | 4 |  | 83 | 92 | 33 | 17 |  |  |  | 50 |  |
| Wei Liu et al. <sup>(15)</sup> | China | 78 | 39 | 39 | 38 |  | 44 |  |  |  |  |  | 26 |  |
| Zhen Li et al. <sup>(38)</sup> | China | 193 | 95 | 98 | 57 | 89 | 69 | 39 | 18 | 36 | 12 | 28 | 28 | 66 |
| Tao Chen et al. <sup>(39)</sup> | China | 274 | 171 | 103 | 62 | 91 | 68 | 50 | 28 | 44 | 44 | 11 | 72 | 41 |
| Bicheng Zhang et al. <sup>(40)</sup> | China | 82 | 54 | 28 | 72.5 | 78 | 65 | 46 | 12 | 63 |  |  | 100 |  |
| Shaobo Shi et al. <sup>(6)</sup> | China | 416 | 205 | 211 | 64 | 80 | 35 | 13 | 4 | 28 |  | 2 | 23 | 14 |
| Luwen Wang et al. <sup>(41)</sup> | China | 116 | 67 | 49 | 54 |  |  |  |  |  |  |  | 10 |  |
| Matt Arentz et al. <sup>(42)</sup> | USA | 21 | 11 |  |  | 52 | 48 |  |  | 76 |  | 2 |  | 52 |
| TieLong Chen et al. <sup>(43)</sup> | China | 203 | 108 | 95 | 54 | 89 | 60 | 8 |  | 2 |  |  |  | 13 |
| Guqin Zhang et al. <sup>(44)</sup> | China | 221 | 108 | 113 | 55 | 91 | 61 | 71 | 11 | 29 | 8 | 5 | 22 | 5 |
| Jianlei Cao et al. <sup>(45)</sup> | China | 102 | 53 | 49 | 54 | 81 | 49 | 55 | 11 | 34 | 15 | 20 | 20 | 17 |
| Ling Hu et al. <sup>(46)</sup> | China | 323 | 166 | 157 | 61 | 84 | 51 |  |  | 4 | 7 | 5 | 4 |  |
| Chaolin Huang et al. <sup>(22)</sup> | China | 41 | 30 | 11 | 49 | 98 | 76 | 44 | 3 | 55 | 12 | 0.7 | 29 | 15 |
| Xiaochen Li et al. <sup>(47)</sup> | China | 548 | 279 | 269 | 60 | 95 | 76 | 47 | 33 | 57 | 22 | 17 | 38 | 17 |
| Jiangshan Lian et al. <sup>(48)</sup> | China | 136 | 58 | 78 |  | 85 | 63 | 18 |  | 13 |  | 2 | 17 | 0 |
| Jiangshan Lian et al. <sup>(48)</sup> | China | 652 | 349 | 303 |  | 80 | 65 | 18 |  | 3 |  | 2 | 5 | 0 |
| Suxin Wan et al. <sup>(49)</sup> | China | 135 | 72 | 63 | 47 | 89 | 77 | 33 | 13 | 13 | 7 | 4 | 16 | 0.7 |
| Qing Deng et al. <sup>(50)</sup> | China | 112 | 57 | 55 | 65 | 88 | 71 |  |  | 56 | 13 |  |  | 0 |
| Fei Shao et al. <sup>(51)</sup> | China | 136 | 90 | 46 |  | 38 | 52 | 49 | 20 | 75 |  |  |  | 0 |
| Wen-Jun Tu et al. <sup>(52)</sup> | China | 25 | 19 | 6 | 70 |  |  |  |  |  | 72 | 76 | 92 | 100 |
| Wen-Jun Tu et al. <sup>(52)</sup> | China | 149 | 60 | 89 | 51 |  |  |  |  |  | 5 | 5 | 5 | 0 |
| Fan Yang et al. <sup>(53)</sup> | China | 91 | 49 | 43 |  |  |  |  |  |  | 34 | 17 | 80 | 100 |
| Qingchun Yao et al. <sup>(54)</sup> | China | 108 | 43 | 65 | 52 | 74 | 78 | 26 | 8 | 14 | 7 | 15 | 42 | 11 |

\*Abbreviations: No=Number; P=prevalence.

**Fig 1.**
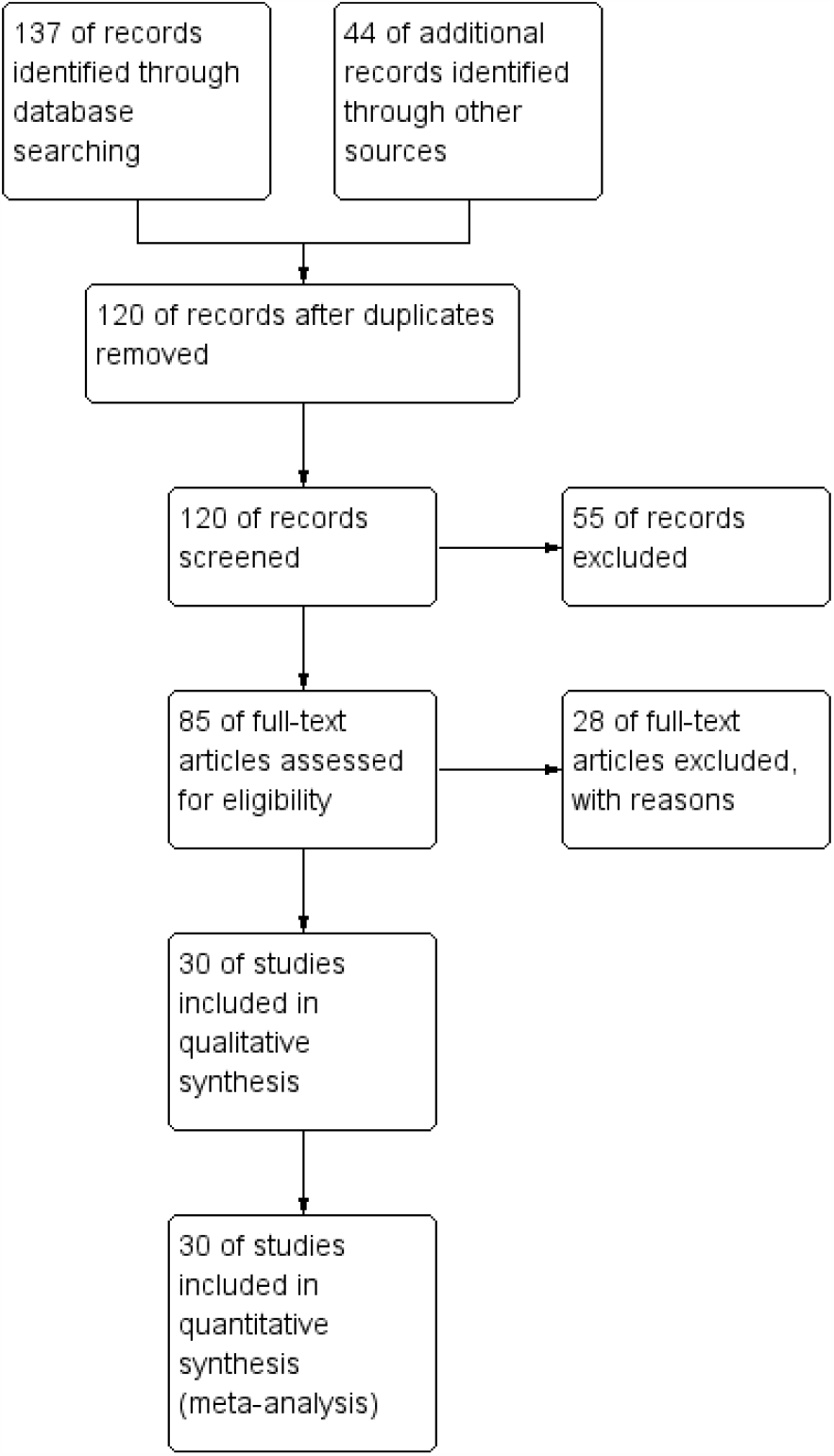
Study flow diagram.

### 3.2. Demographic Characteristics of included study

Pooling all extracted data together, the total number of COVID-19 positive patients was 6 389 including 1402 patients with ARDS, 494 patients with acute cardiac injury and 418 patients with AKI. Selected studies have been conducted primarily in China except 1 in the United States. According to the average calculated from the articles reporting the median age of subjects, the mean age of patients were 57.64 and 45.29%, (95% CI: 42.61% to 47.99%; I^2^=74.56%) of the patients were female.

### 3.3. Clinical Manifestations

Our meta-analysis revealed that the most common symptoms of COVID-19 were fever 84.30% (95% CI: 77.13-90.37; I^2^=97.74%), cough 63.01% (95% CI: 57.63-68.23; I^2^=93.73%), dyspnea 37.16% (95% CI: 27.31-47.57%; I^2^=98.32%), fatigue 34.22% (95% CI: 26.29-42.62; I^2^=97.29%) and diarrhea 11.47% (95% CI: 6.96-16.87; I^2^=95.58%), respectively.

### 3.4. Complications

Cardiac and renal complications in these patients were also assessed. These most common respiratory complication was ARDS 33.15% (95% CI: 23.35-43.73; I^2^=98.56%). The prevalence of arrhythmia, acute cardiac injury and heart failure were 16.64% (95% CI: 9.34-25.5; I^2^=92.29%), 15.68% (95% CI: 11.1-20.97; I^2^=92.45%), and 11.50% (95% CI: 3.45-22.83; I^2^=89.48%), respectively. Unfortunately, few related articles were available for analyzing the prevalence of arrhythmia and heart failure. Furthermore, the primary analysis of AKI showed a prevalence of 9.87% (95% CI: 6.18-14.25; I^2^=95.64%) (Table 2).

**Table 2.** Statistical analysis of reviewed studies.

|  | Number of study | Prevalence (%) | 95% CI | I<sup>2</sup> (%) | P Value for heterogeneity |
| --- | --- | --- | --- | --- | --- |
| **Mortality** | 22 | 12.29 | (6.20-19.99) | 98.29 | 0.00 |
| Mortality in China | 21 | 11.20 | (5.36-18.75) | 98.34 | 0.00 |
| Mortality in USA | 1 | 52.38 | (29.78-74.29) |  |  |
| **Gender** |  |  |  |  |  |
| Female | 29 | 45.29 | (42.61-47.99) | 74.56 | 0.00 |
| Male | 29 | 54.72 | (52.04-57.39) | 74.33 | 0.00 |
| **Clinical Manifestations** |  |  |  |  |  |
| Fever | 24 | 84.30 | (77.13-90.37) | 97.74 | 0.00 |
| Cough | 25 | 63.01 | (57.63-68.23) | 93.73 | 0.00 |
| Dyspnea | 23 | 37.16 | (27.31-47.57) | 98.32 | 0.00 |
| Fatigue | 21 | 34.22 | (26.29-42.62) | 97.29 | 0.00 |
| Diarrhea | 18 | 11.47 | (6.96-16.87) | 95.58 | 0.00 |
| **Complications** |  |  |  |  |  |
| <u>Respiratory</u> |  |  |  |  |  |
| ARDS | 27 | 33.15 | (23.35-43.73) | 98.56 | 0.00 |
| <u>Cardiovascular</u> |  |  |  |  |  |
| Acute Cardiac injury | 15 | 13.77 | (9.66-18.45) | 91.36 | 0.00 |
| Arrhythmia | 5 | 16.64 | (9.34-25.50) | 92.29 | 0.00 |
| Heart failure | 4 | 11.50 | (3.45-22.83) | 89.48 | 0.00 |
| <u>Renal</u> |  |  |  |  |  |
| AKI | 21 | 8.40 | (5.15-12.31) | 95.22 | 0.00 |

### 3.5. Outcomes

Based on the primary analysis, the mortality rate was 21.70% (95% CI: 12.39-32.70; I^2^=98.81%), but after omitting 3 of our included studies conducted only on deceased patients, the total mortality rate was calculated 12.29% (95% CI: 6.20-19.99; I^2^=98.29%). We also conducted a subgroup analysis to determine the mortality rate by location. As stated above, our articles were mainly from China with a mortality rate of 11.20 (95% CI: 5.36-18.75; I^2^=98.34%) and one of these was from the USA with a mortality rate of 52.38 (95% CI: 29.78-74.29).

### 3.6. Sensitivity Analysis

According to the primary analysis, the prevalence of acute cardiac injury was 15.68% (95% CI: 11.1-20.97; I^2^=92.45%), but after omitting a part of the data from the Wen-Jun Tu study and conducting sensitivity analysis (omitting outlier data) the prevalence was 13.77% (95% CI: 9.66-18.45; I^2^=91.36%) (Figure 2). Apparently, the primary analysis of AKI showed a prevalence of 9.87% (95% CI: 6.18-14.25; I^2^=95.64%), and after omitting a part of the data from Wen-Jun Tu study and conducting sensitivity analysis (removed outlier data) the prevalence was 8.40% (95% CI: 5.15-12.31; I^2^=95.22%) (Figure 3), respectively.

**Fig 2.**
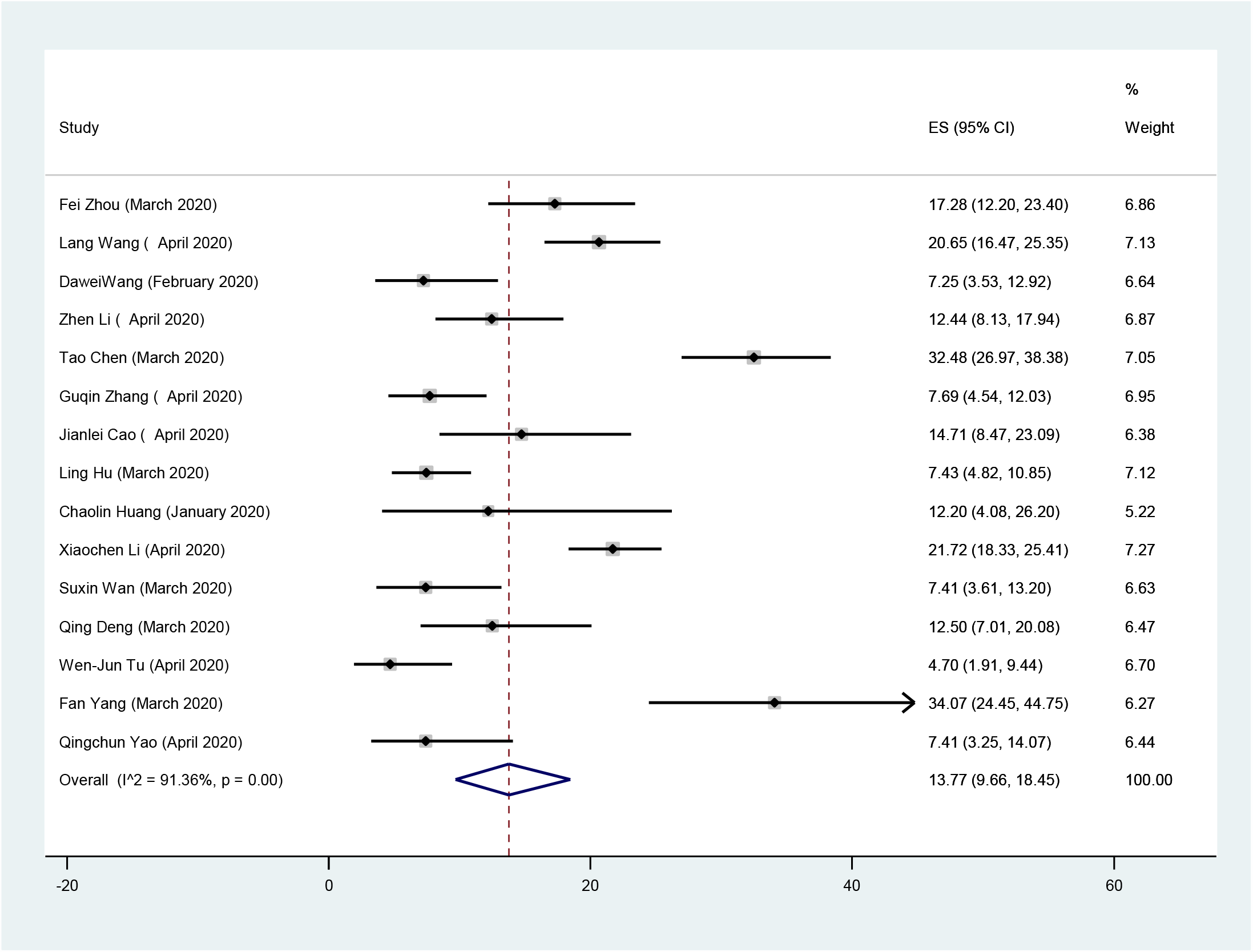
Forest Plot of the prevalence of acute cardiac injury in COVID-19 patients. Each square shows effect estimate of individual studies with their 95% CI. Size of squares is proportional to the weight of each study in the meta-analysis. In this plot, studies are shown in the order of publication date and first author’s names (based on a random effects model).

**Fig 3.**
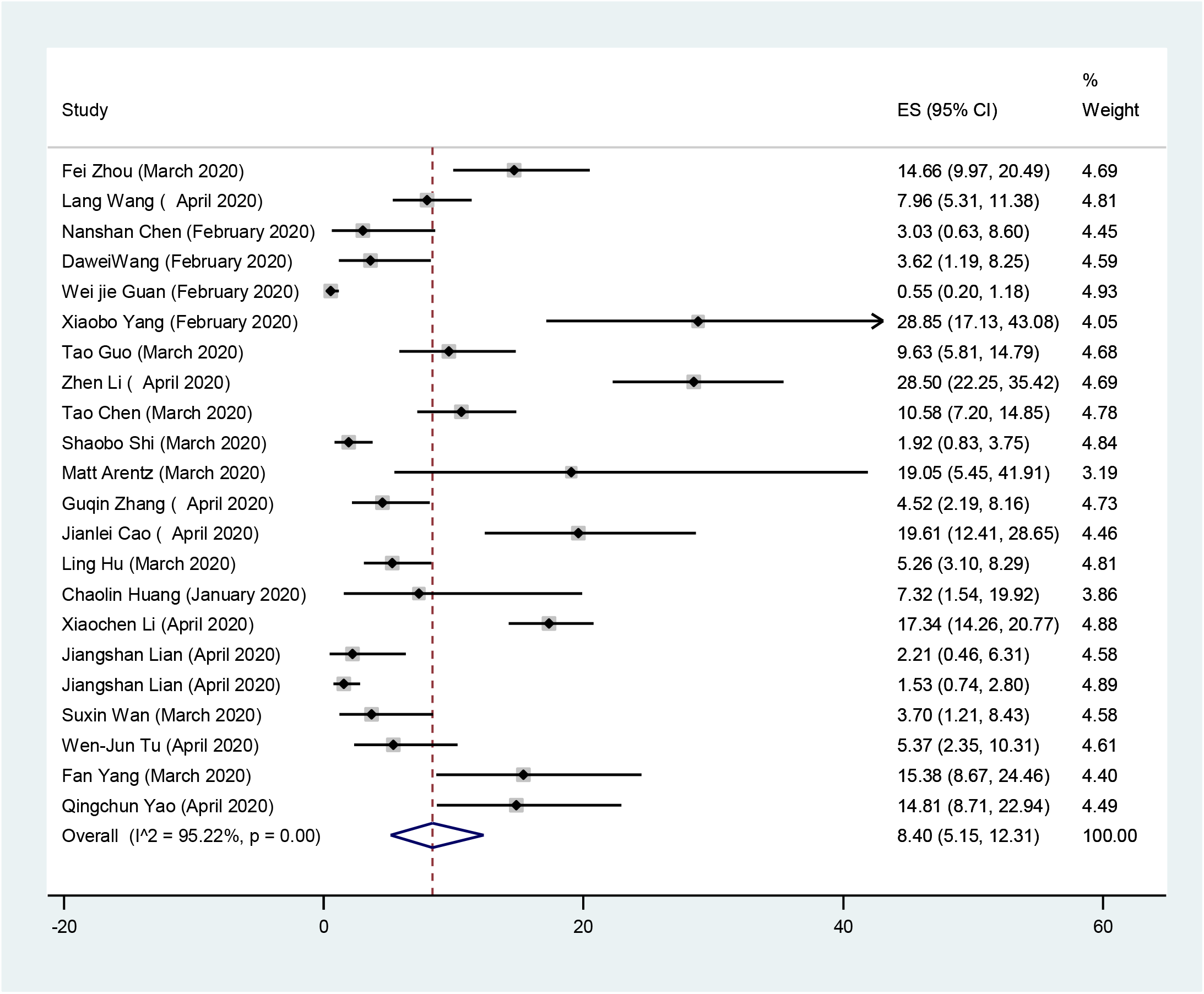
Forest Plot of the prevalence of acute kidney injury (AKI) in COVID-19 patients. Each square shows effect estimate of individual studies with their 95% CI. Size of squares is proportional to the weight of each study in the meta-analysis. In this plot, studies are shown in the order of publication date and first author’s names (based on a random effects model).

### 3.7. Publication Bias

Figure 4 shows the Beggs funnel plot of studies related to cardiac and renal injury in COVID-19 patients. The interpretation of this plot showed no sign of publication bias in these studies (p=0.08); this result implies that reports have been published with both positive and negative results (Figure 4).

**Fig 4.**
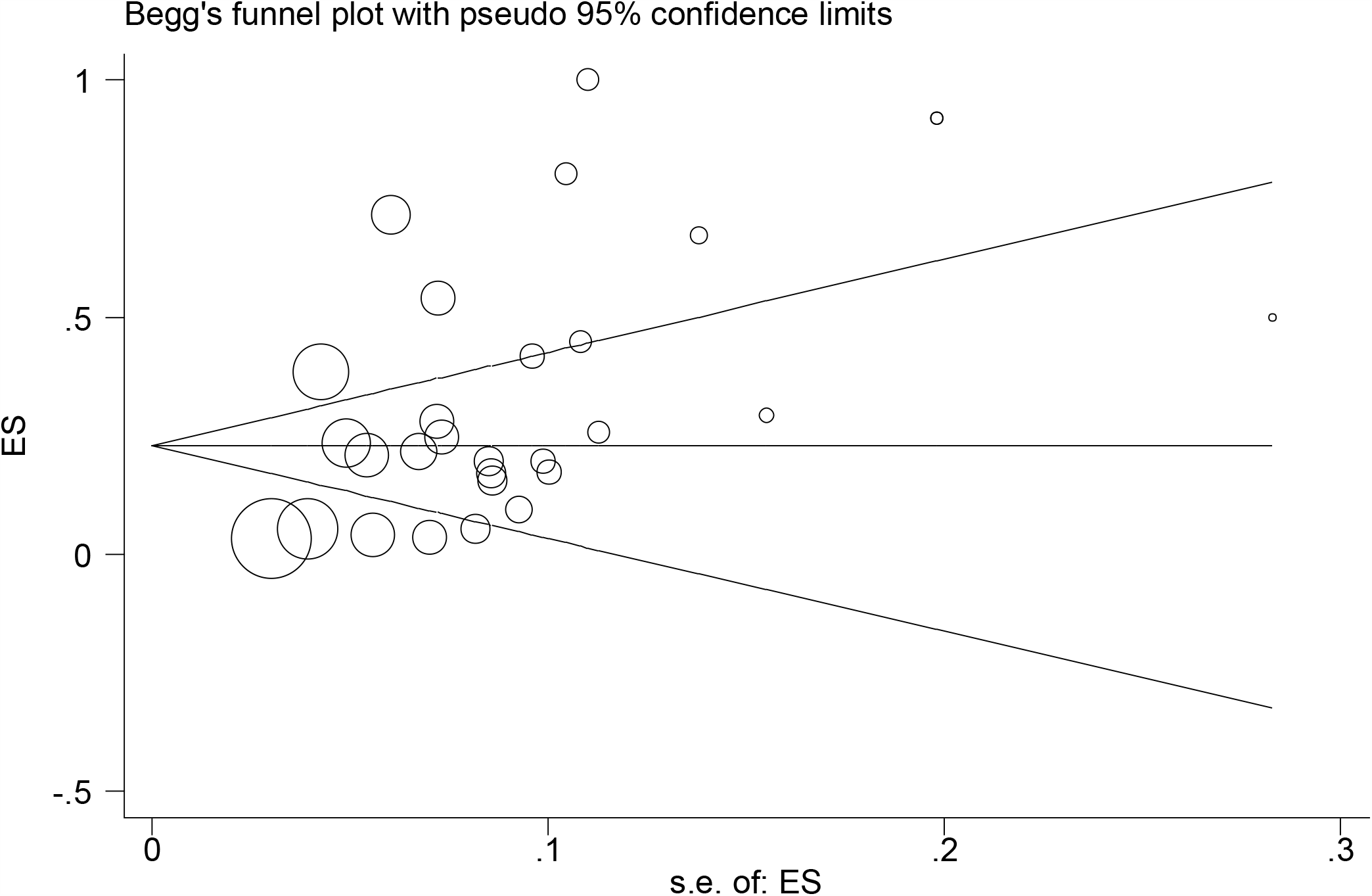
Begg’s funnel plot for publication bias.

## 4. Discussion

From last days of 2019, a novel coronavirus outbreak has emerged, which has been further named coronavirus disease 2019 (COVID-19), and has been spreading around the world since then (10). The city of Wuhan in China was the first infected area and the starting point of this pandemic (11). COVID-19 is highly transmittable and has a great ability to cause to the cluster of cases (12, 13). To date, no definite treatment has been discovered for COVID-19. Current recommendations regarding this disease include preventive measures to reduce the risk of infection (14, 15). Unfortunately, in severe cases of COVID-19, the disease can lead to acute respiratory distress syndrome (ARDS), which is one of the deadliest complications of COVID-19 and can contribute to death. According to Tian et al (16), the prevalence of severe and mild outcomes were 18% of severe cases, while other 82% were common cases (including: including mild cases (73.3%), non-pneumonia cases (4.2%) and asymptomatic cases (5.0%), respectively).

Our meta-analysis is conducted on data extracted from 30 published studies. These studies collected data from laboratory-confirmed COVID-19 patients and were mainly conducted in Chinese hospitals. According to previous reports during SARS-CoV and MERS-CoV outbreaks, these corona viruses have been seen to affect males more than females (17, 18). And also regarding COVID-19, Yang et al revealed that males are at greater risk than females (19). Our results also confirm these literatures; SARS-CoV-2 has a higher risk in men (54.72% (95% CI: 52.04-57.39; I^2^=74.33%)) compared to women (45.29% (95% CI: 42.61-47.99; I^2^=74.56%)). This difference could be the result of stronger innate immune responses in women (20), more harmful lifestyle habits (such as smoking) in men (21) and also greater exposure to occupational hazards in men (e.g. Hunan wet market in China) (22). According to the age-based analysis, the mean age of subjects were 57.64, which is close to the number reported in previous studies (23). Elderly patients have indeed been shown to be at greater risk of more serious complications and further death, which may be related to higher prevalence of comorbidities between elderlies (24) or their weakened immune system (21).

We also extracted data about most frequent symptoms of COVID-19. According to our analysis, fever 84.30% (95% CI: 77.13-90.37; I^2^=97.74%), cough 63.01% (95% CI: 57.63-68.23; I^2^=93.73%), dyspnea 37.16% (95% CI: 27.31-47.57%; I^2^=98.32%), fatigue 34.22% (95% CI: 26.29-42.62; I^2^=97.29%) and diarrhea 11.47% (95% CI: 6.96-16.87; I^2^=95.58%) are the most prevalent symptoms of COVID-19, respectively. These result are almost similar to previous reports (19, 23).

As we looked at the included studies, we realized that the most reported complication was ARDS; that its prevalence was 33.15% (95% CI: 23.35-43.73; I^2^=98.56%). ARDS occurs as a result of the accumulation of fluids in alveoli, and this fluid prevents the lungs from receiving enough air. The cause of this phenomenon is the leakage of fluid from capillaries into alveoli, due to damage of their walls (25). This high prevalence of ARDS is justifiable due to the large pneumonia caused by this virus. From other non-respiratory outcomes, acute cardiac injury and AKI have been the most reported. Acute myocardial injury and its complications were observed in 9.5% of all terminally ill COVID-19 patients in Italy (until 13 April 2020) (26). According to our analyses, the prevalence of acute cardiac injury was 13.77% (95% CI: 9.66% to 18.45%; I^2^=91.36%). This complication has signs and symptoms similar to COVID-19 respiratory complications and may develop at any stage of this disease.

Acute myocardial injuries in patients with COVID-19 include arrhythmias, heart failure, cardiac arrest, acute coronary syndromes, cardiomyopathy, myocarditis, cardiogenic shock, pericarditis and pericardial effusion (27). In our analysis, the prevalence of arrhythmia 16.64% (95% CI: 9.34-25.5; I^2^=92.29%) and heart failure 11.50% (95% CI: 3.45-22.83; I^2^=89.48%) were assessed as well, but due to insufficiency of number of articles and lack of enough data, they actually may overestimate the real numbers. Symptoms related to acute cardiac injury are dyspnea, severe fatigue, chest pain and palpitation. Markers that can help diagnose an acute myocardial injury are high sensitivity troponin I (hs-cTnI) or T (hs-cTnT) and NT-proBNP. Performing an electrocardiogram (ECG) can also be helpful (ECG changes show myocardial ischaemia). Troponin level also has diagnostic value, as inflammatory responses of heart to severe illness can lead to elevated troponin levels (27). The prevalence of AKI was 8.40% (95% CI: 5.15-12.31; I^2^=95.22%). AKI is described as a sudden failure in kidney function, which includes both structural damage (injury) and loss of function (failure). Markers that can help diagnose AKI are serum creatinine (sCr) and/or urine output (UO) (28).

As stated before, the main focus of this study is to determine the most frequent respiratory and non-respiratory complications in COVID-19 patients, which can help find the causes of worst outcomes and death in these patients. On March 3, 2020, the mortality rate of COVID-19 was reported by 3.4% by World Health Organization (WHO) (29). While according to our analysis, the mortality rate was 12.29% (95% CI: 6.20-19.99; I^2^=98.29%) (this number was calculated after omitting the studies that included only deceased patients). It should be noted that most of our included articles were conducted on severe cases, so this higher mortality rate is reasonable. We also performed a subgroup analysis to determine the mortality rate based on the location, which the results was 11.20% (95% CI: 5.36-18.75; I^2^=98.34%) in China and 52.38% (95% CI: 29.78-74.29) in the USA. These results have also overestimated the mortality rate of patients with COVID-19, both due to the inclusion of predominantly severe cases and the lack of sufficient studies with useful data. Since no systematic review and meta-analysis on the non-respiratory outcomes of COVID-19 have yet been published, we look forward this article having useful results for the medical society.

## 5. Conclusion

COVID-19 infection has a high morbidity rate especially in elderly patients. These severe patients require supportive medical care, which can place a huge burden on nations. There are several complications that can put patients in critical conditions. The most frequent clinical outcome of COVID-19 is respiratory complications, in particular ARDS. There are also some non-respiratory outcomes in COVID-19 patients. Although these outcomes are less common than respiratory outcomes, they can have fatal effects on patients. Two of the most non-respiratory complications in COVID-19 patients are acute cardiac injury and AKI. Knowing this information gives clinicians a brighter insight to what they are fighting against. Since these data are updating day by day, further evaluation of these clinical outcomes is recommended.

## Data Availability

Available.

## 6. Acknowledgment

The authors are grateful to Shahid Beheshti University of Medical Sciences, Tehran, IR Iran for their collaborative efforts.

